# An International Review of Tobacco Use and the COVID-19 Pandemic: Examining Hospitalization, Asymptomatic Cases, and Severity

**DOI:** 10.1101/2020.06.12.20129478

**Authors:** Lachezar L. Balabanski

## Abstract

**Background and objectives:** Since the World Health Organization (WHO) declared a public health emergency of imminent concern in March 2020, the novel coronavirus (SARS-CoV-2) and its related disease (COVID-19) has become a topic of much needed research. This study primarily focused on what effect smoking had on hospitalization; however, asymptomaticity, and severity were discussed.

**Data:** Data was collected through searches on databases including PubMed and Google Scholar. Eligibility criteria included being RT-PCR verified and including smoking data.

**Discussion:** This study found that smokers were significantly underrepresented in COVID-19 hospitalization on a purely epidemiological level in some areas, including China and Manhattan, but not others: Seattle, Greater New York City Area, and Italy. Furthermore, smokers were equally represented in asymptomatic populations, but smokers will likely experience a more severe manifestation of the disease if they are symptomatic. Further inquiry into possible mechanisms by which the observed epidemiological effect is necessary, as it has implications for recommendations on loosening restrictions on social distancing measures.

**Conclusions and Recommendations:** This study recommends that smokers consider themselves to be at higher risk for COVID-19, as they may experience a more severe manifestation of the disease. Health care providers and policy makers should consider smokers at higher risk as well, as there is evidence to claim that smokers may contract a more severe form of COVID-19.

## INTRODUCTION

The SARS-CoV-2 virus originated in late December in Wuhan, China, where it has since spread to many parts of the world.[1, 2] As of 9 May 2020, an estimated 3.9 million people worldwide have been diagnosed with the SARS-CoV-2 virus. Clearly, the disease is continuing to spread, as according to the WHO, about 95,000 cases have developed in the past day[3]; virtually every country, at this point, has had some exposure to the virus. The virus is disproportionately severe in the elderly and those with comorbidities,[4,5] prompting many countries to undergo self-isolation procedures.[6]

Smoking has been implicated as a risk factor for countless viral infections, including influenza.[7] Thus, smoking has been hypothesized to be a risk factor for COVID-19, especially because the disease is respiratory.

It is now well known that SARS-CoV-2 uses the Angiotensin-converting enzyme 2 (ACE2) receptor for entry into a target cell with greater affinity than did SARS-CoV.[8] Furthermore, the ACE2 receptor is upregulated in those who smoke.[9] Thus, it has been suggested that those who smoke are more susceptible to SARS-CoV-2, and limited, anecdotal evidence has been presented to support this claim.[10, 11] However, very few of the studies used statistical tests, and, when they were used, very few yielded significance, and virtually no studies examined the disease and tobacco use outside of China. There is some limited evidence published to argue that there is no association between smoking and hospitalization due to or severity of COVID-19.[12, 13] However, similar to the previous studies, they did not examine data from outside China.

International analyses are very important regarding the COVID-19 pandemic. Ethnic and genetic disparities in expression of the *ACE2* gene could possibly explain the discrepancy in case-fatality rate between China and Italy, for example.[14] Furthermore, discrepancies in gender between those who smoke and do not smoke are very significant in many countries,[15] and has been implicated as a possible explanation for the discrepancy in male-female fatality due to COVID-19.[16] Lastly, air quality has been correlated with COVID-19 severity by country; however, it remains unclear whether this relationship is causal.[17] Thus studying the two variables (hospitalization due to COVID-19 and smoking rates) between nations will increase the likelihood that the correlation observed is strictly due to smoking.

Many studies have been published regarding COVID-19 and the association with smoking; however, this is the first study, to the knowledge of the author, detailing asymptomaticity of COVID-19 in smokers and that reviewed multiple centers of the pandemic. This systematic review aims to examine whether or not those hospitalized for COVID-19 were disproportionately smokers. It was hypothesized that there would be no correlation between the two variables. In other words, smokers were hypothesized to be proportionally represented in hospitalizations.

## METHODS

Firstly, data was acquired by search query for, “clinical characteristics of COVID-19,”, “SARS-CoV-2 clincal features,” or for “[SARS-COV-2 OR COVID-19] AND [smoking OR tobacco]” and related terms through the databases PubMed, ScienceDirect, and Google Scholar. No language restrictions were placed on the studies. Inclusion criteria were as follows: full text was available, study was subject to peer-review, all diagnoses were verified by RT-PCR, and smoking data were provided. The search was conducted from 30 April 2020 to 9 May 2020 Of 5,360 potential results, only 6 met the eligibility criteria. Data was extracted from data tables provided by the study authors in the article text. No data was pooled or combined with other studies’ data. The data presented was assumed to be free of bias, as all diagnoses were RT-PCR verified and studies included all individuals who were hospitalized for the disease for some period. Individuals who were former smokers were counted as smokers, whereas those who had never smoked were counted as non-smokers for a more conservative analysis. Based on data released by governments of the various nations studied, the proportion of active smokers in the general population was used. For country-level studies, the published 2015 WHO[15] smoking data was used; however, in the case of United States centered studies, smoking data was collected from state government reporting.[26, 27] All the data found above can be found in Table I.

**Table I:** Smoking and Geographic Data

| **Author(s)** | **Geographic Location** | **Smokers (count)** | **Non-smokers (count)** | **Total** | **Expected smoking proportion** |
| --- | --- | --- | --- | --- | --- |
| Guan et al. [20] | China | 158 | 927 | 1085 | 25.3% [15] |
| Hu et al.<sup>a</sup> [21] | China | 2 | 17 | 19 | 25.3% [15] |
| Bhatraju et al. [22] | Seattle | 5 | 19 | 24 | 13.5% [27] |
| Goyal et al. [23] | Manhattan, NYC | 20 | 373 | 393 | 9.60% [26] |
| Goyal et al. (Requiring Mechanical Ventilation) [23] | Manhattan, NYC | 6 | 124 | 130 | 9.60% [26] |
| Richardson et al. [24] | Greater NYC Area | 2133 | 3567 | 5700 | 10.90% [26] |
| Inciardi et al. [25] | Italy | 17 | 82 | 99 | 24.0% [15] |

<sup>a</sup> The Hu et al. data included asymptomatic patients only

Next, a 1-tailed, 1-sample Pearson’s chi-squared test was conducted for each of the relevant studies’ data with significance of 0.05. It was alternatively hypothesized that hospitalization and smoking status would not be completely independent. For studies that had fewer than 30 patients, a Fisher Exact test was used with the same hypotheses and level of significance. Yule’s Q were calculated for all groups to demonstrate directionality and strength of the correlation. In addition, an odds ratio was calculated for all studies. All statistical tests were performed using Microsoft Excel. The relevant statistics and p-values are presented in Table II.

**Table II:** Summary Table of Statistical Testing

| **Author** | **$\chi^2$-statistic** | **p-value** | **Yule's Q** |
| --- | --- | --- | --- |
| Guan et al.[20] | 66.19 | $p < 0.0001$ | -0.330 |
| Hu et al.[21] | N/A<sup>b</sup> | $p = 0.405$ | -0.484 |
| Bhatraju et al.[22] | N/A<sup>b</sup> | $p = 0.701$ | 0.255 |
| Goyal et al. [23] | 9.21 | $p = 0.002$ | -0.329 |
| Goyal et al. (Requiring Mechanical Ventilation) [23] | 3.72 | $p = 0.054$ | -0.374 |
| Richardson et al.[24] | 1093.09 | $p < 0.0001$ | 0.660 |
| Inciardi et al.[25] | 2.53 | $p = 0.111$ | -0.207 |

<sup>b</sup> Sample size was too small to use the chi-squared test, Fisher Exact test was used instead, as described above.

## RESULTS

A total of 7,320 patients were included in this study, of whom 2,335 (31.9%) had some prior history of smoking. About 15.1% of those cases were hospitalized in China and 83.6% hospitalized in the United States; the remaining approximate 1.4% were hospitalized in Italy. Table I contains general information about the data and studies, and Table II contains results from statistical testing applied on those data in Table I. After reading, all studies examined were free from bias that would significantly alter the conclusions drawn.

Yule’s Q is a measure of the strength of correlation for the two variables (smoking and hospitalization). The more extreme the Q value is, the larger the correlation is (ranging from -1 to 1). Here, a negative Q value indicates an inverse correlation between smoking and hospitalization, and a positive Q value indicates a positive, direct correlation between smoking and hospitalization.

## DISCUSSION

Based on the results in Table II, in China, smoking appears to have a preventative effect on hospitalization for COVID-19 (χ^2^ = 66.19, p < 0.0001, Q = -0.33), at least on the epidemiological level. These measurements were extremely significant with a large sample size (n = 1085). This result is consistent with several similar findings that only examined patient data from China and East Asia.[12,13]

However, when examining geographic regions outside of China, the data appears to be a lot more mixed. Seattle and Italy, with relatively smaller sample sizes (n = 24 and n =99) both did not reach significance (p = 0.701 and χ^2^ = 2.53; p = 0.111, respectively), meaning smokers were equally represented in the hospitalized patients in those areas. Manhattan, New York was the only epicenter outside of China to reach statistical significance for a preventative effect (χ^2^ = 9.21, p = 0.002, Q = -0.329). And this is possibly contradicted by the larger, more significant finding in the greater New York City Area, that smokers were more likely to be hospitalized for COVID-19 (χ^2^ = 1093.09, p < 0.0001, Q = 0.660). Interestingly, New York City was one of the areas in the United States with the poorest air quality in terms of NO_2_ concentration.[17]

By using the requirement of mechanical ventilation as an assay for severity, there is evidence to suggest that smokers get a more severe manifestation of the disease. Smokers were underrepresented in the population of hospitalized patients in Manhattan (χ^2^ = 9.21, p = 0.002, Q = -0.329), but those who needed mechanical ventilation were proportionately smokers (χ^2^ = 3.72, p = 0.054, Q = -0.374). This suggests that smokers either contracted a more severe form of the disease or were more susceptible to severe infection by the otherwise less severe disease. These findings are consistent with those of Patanavanich & Glantz.[18]

These data may suggest that, when adjusted for air quality, there is a direct association with the severity of the disease and tobacco use, but not between hospitalization and tobacco use. Further research should focus on the mechanisms by which this could possibly occur.

Lastly, the relationship between asymptomatic patients and their tobacco use was examined. The data indicated that smokers were proportionally represented in asymptomatic patients. The relatively small sample size could be the reason for the lack of significance; however, based on the available evidence, there is reason to claim that smokers are equally represented in the population of asymptomatic patients. Asymptomatic patients are important to study with respect to smokers, because they could be responsible for up to 79% of the documented, symptomatic cases.[19]

Beyond risk factors analysis, this study has implications in treatment for COVID-19. Research done by Gonzalez-Rubio et al.[13] suggests nicotine has potential for treatment of COVID-19 by suppressing cytokine storm.

Because this study was conducted on an epidemiological level, there are many possible confounds that will necessarily dilute the findings of the study. As Berlin, Thomas, Le Faou and Cornuz[10] observed, comorbidities, including hypertension, obesity, coronary heart disease, etc., are inherently inseparable from epidemiological review, such as the one conducted in this study. Furthermore, several countries’ tobacco use significantly differs by gender.[15] Thus, the underrepresentation of smokers could be correlated somehow with gender distribution. A similar sentiment was detailed by Cai.[16] The results of this study suggest that tobacco smokers should undergo similar isolation procedures to non-smokers; however, in areas with poorer air quality, smokers should err on the side of caution and consider themselves at higher risk for COVID-19.

Review bias has been attempted to be minimized by this study. By doing a systematic review, review bias was minimized. Some concerns for overreporting of cases have come up in the literature recently.[28] By only including diagnoses verified by RT-PCR, overreporting of COVID-19 cases was also minimized.

This study examined data between nations and in 4 major epicenters of the pandemic. Aggregating data from other epicenters of the pandemic, especially from Europe, the Middle East, Africa, and South America, would be very beneficial. Studies similar to this one are critical at this stage of the pandemic, especially in assessing the risks of various recommendations for which countries, states, or municipalities can ease prevention measures.

For future research purposes, databases recording smoking incidence in COVID-19 should be published to address concerns of current smokers or those related to smokers, as well as to fully identify all the potential risk factors of the disease. The more understanding of the risk factors of the disease, the better the understanding of the mechanism by which it acts is understood. Furthermore, a richer understanding of the biological processes which take place in smokers who are hospitalized with COVID-19 would be invaluable to research efforts to stop the pandemic.

## CONCLUSION

Based on the evidence presented here, it is sufficient to say that the exact variables influencing COVID-19 hospitalizations remain unclear. There are multiple epicenters where smokers are underrepresented in hospitalizations due to COVID-19. Despite this, smokers should act cautiously, because they may be at higher risk depending on the air quality of their geographic location.

## Data Availability

All data is presented in the manuscript itself. If necessary, it can be given upon a reasonable request (with analysis) in Excel format.

## REFERENCES

1. WHO. WHO Director-General’s opening remarks at the media briefing on COVID-19 -11 March 2020. Published 2020. Accessed May 7, 2020. https://www.who.int/dg/speeches/detail/who-director-general-s-opening-remarks-at-the-media-briefing-on-covid-1911-march-2020

2. Tang X, Wu C, Li X, et al. On the origin and continuing evolution of SARS-CoV-2. Published online 2020. doi:10.1093/nsr/nwaa036/5775463

3. WHO. COVID-19 situation reports. Published 2020. Accessed May 9, 2020. https://www.who.int/emergencies/diseases/novel-coronavirus-2019/situation-reports/

4. Liu K, Chen Y, Lin R, Han K. Clinical features of COVID-19 in elderly patients: A comparison with young and middle-aged patients. J Infect. Published online March 27, 2020. doi:10.1016/j.jinf.2020.03.005

5. Yang J, Zheng Y, Gou X, et al. Prevalence of comorbidities and its effects in coronavirus disease 2019 patients: A systematic review and meta-analysis. Int J Infect Dis. 2020;94:91–95. doi:10.1016/j.ijid.2020.03.017

6. Musinguzi G, Asamoah BO. The Science of Social Distancing and Total Lock Down: Does it Work? Whom does it Benefit? Electron J Gen Med. 2020;2020(6):2516–3507. doi:10.29333/ejgm/7895

7. Lawrence H, Hunter A, Murray R, Lim WS, McKeever T. Cigarette smoking and the occurrence of influenza – Systematic review. J Infect. 2019;79(5):401–406. doi:10.1016/j.jinf.2019.08.014

8. Zhang H, Penninger JM, Li Y, Zhong N, Slutsky AS. Angiotensin-converting enzyme 2 (ACE2) as a SARS-CoV-2 receptor: molecular mechanisms and potential therapeutic target. Intensive Care Med. 2020;46(4):586–590. doi:10.1007/s00134-020-05985-9

9. Brake SJ, Barnsley K, Lu W, McAlinden KD, Eapen MS, Sohal SS. Smoking Upregulates Angiotensin-Converting Enzyme-2 Receptor: A Potential Adhesion Site for Novel Coronavirus SARS-CoV-2 (Covid-19). J Clin Med. 2020;9(3):841. doi:10.3390/jcm9030841

10. Berlin I, Thomas D, Le Faou A-L, Cornuz J. COVID-19 and smoking. Published online 2020. doi:10.1093/ntr/ntaa059/5815378

11. Vardavas CI, Nikitara K. COVID-19 and smoking: A systematic review of the evidence. Tob Induc Dis. 2020;18. doi:10.18332/TID/119324

12. Lippi G, Henry BM. Active smoking is not associated with severity of coronavirus disease 2019 (COVID-19). Eur J Intern Med. 2020;75:107–108. doi:10.1016/j.ejim.2020.03.014

13. Gonzalez-Rubio, J., Navarro, C., Lopez-Najera, E., Lopez-Najera, A., Jimenez-Diaz, L., Navarro López, J. D. PhD, & Najera A. Cytokine release syndrome (CRS) and nicotine in COVID-19 patients: trying to calm the storm. Published online 2020. doi:10.31219/osf.io/chd2k

14. Asselta R, Paraboschi EM, Mantovani A, Duga S. ACE2 and TMPRSS2 Variants and Expression as Candidates to Sex and Country Differences in COVID-19 Severity in Italy. SSRN Electron J. Published online April 2, 2020. doi:10.2139/ssrn.3559608

15. WHO. GHO | By category | Tobacco use - Data by country. Published 2015. Accessed May 9, 2020. http://apps.who.int/gho/data/node.main.651.

16. Cai H. Sex difference and smoking predisposition in patients with COVID-19. Lancet Respir Med. 2020;8(4):e20. doi:10.1016/S2213-2600(20)30117-X

17. Pansini R, Fornacca D. COVID-19 higher induced mortality in Chinese regions with lower air quality. doi:10.1101/2020.04.04.20053595

18. Patanavanich R, Glantz SA. Smoking is Associated with COVID-19 Progression: A Meta-Analysis. medRxiv. Published online April 16, 2020:2020.04.13.20063669. doi:10.1101/2020.04.13.20063669

19. Li R, Pei S, Chen B, et al. Substantial undocumented infection facilitates the rapid dissemination of novel coronavirus (SARS-CoV2). Science. Published online March 16, 2020. doi:10.1126/science.abb3221

20. Guan W, Ni Z, Hu Y, et al. Clinical Characteristics of Coronavirus Disease 2019 in China. N Engl J Med. 2020;382(18):1708–1720. doi:10.1056/NEJMoa2002032

21. Hu Z, Song C, Xu C, et al. Clinical characteristics of 24 asymptomatic infections with COVID-19 screened among close contacts in Nanjing, China. Sci China Life Sci. 2020;63(5):706–711. doi:10.1007/s11427-020-1661-4

22. Bhatraju PK, Ghassemieh BJ, Nichols M, et al. Covid-19 in Critically Ill Patients in the Seattle Region — Case Series. N Engl J Med. Published online March 30, 2020. doi:10.1056/nejmoa2004500

23. Goyal P, Choi JJ, Pinheiro LC, et al. Clinical Characteristics of Covid-19 in New York City. N Engl J Med. Published online April 17, 2020. doi:10.1056/nejmc2010419

24. Richardson S, Hirsch JS, Narasimhan M, et al. Presenting Characteristics, Comorbidities, and Outcomes Among 5700 Patients Hospitalized With COVID-19 in the New York City Area. JAMA. Published online April 22, 2020. doi:10.1001/jama.2020.6775

25. Inciardi RM, Adamo M, Lupi L, et al. Characteristics and outcomes of patients hospitalized for COVID-19 and cardiac disease in Northern Italy. doi:10.1093/eurheartj/ehaa388

26. New York Bureau of Tobacco Control. StatShot Vol. 11, No. 4 /May 2018, Prevalence of Current Smoking Among Adults in New York by County, NYS BRFSS 2016.; 2018. Accessed May 7, 2020. https://www.health.ny.gov/prevention/tobacco_control/reports/statshots/

27. Washington State Department of Health. Tobacco Data and Reports. Accessed May 7, 2020. https://www.doh.wa.gov/DataandStatisticalReports/HealthBehaviors/Tobacco

28. Pasquariello P, Stranges S. Excess mortality from COVID-19: a commentary on the Italian experience. doi:10.1007/s00038-020-01399-y

